# Pathogenic desmin variants impair nuclear integrity and drive atrial cardiomyopathy

**DOI:** 10.64898/2026.04.07.26348559

**Authors:** Wei Su, Stan W. van Wijk, Preetam Kishore, Mingxin Huang, Danish Sultan, Leonoor F. J. M. Wijdeveld, Fabries G. Huiskes, Amelie C.T. Collinet, Niels Voigt, Aiste Liutkute, Marijn Brands, Tyler Kirby, Reinier L. van der Palen, Kondababu Kurakula, Kennedy Silva Ramos, Christof Lenz, Ingeborg M Bajema, Karin Y. van Spaendonck-Zwarts, Andreas Brodehl, Hendrik Milting, J. Peter van Tintelen, Bianca J.J.M. Brundel

## Abstract

**Background:** Pathogenic desmin (*DES)* variants have been implicated in early-onset atrial disease, yet the mechanisms by which desmin dysfunction alters atrial structure and function remain unclear. Desmin anchors the cytoskeleton to the nuclear envelope (NE) through the linker of nucleoskeleton and cytoskeleton (LINC) complex, suggesting that defects in this network may drive atrial cardiomyopathy.

**Methods:** Human desmin wild-type (WT) and the pathogenic variants p.S13F, p.N342D, and p.R454W were stably expressed in HL-1 atrial cardiomyocytes. Desmin organization, nuclear morphology, LINC-complex integrity (nesprin-3, lamin A/C), and DNA leakage, assessed by cyclic GMP–AMP synthase (cGAS), were analyzed by confocal microscopy. Action potential duration (APD) and calcium transients (CaT) were measured optically. Human myocardium samples from *DES* variant carriers were analyzed for validation. Data-independent acquisition (DIA) mass spectrometry profiled atrial proteomes from desmin-network (DN) and titin variant carriers and controls. The heat-shock proteins (HSPs) inducer geranylgeranylacetone (GGA) was evaluated for rescue effects.

**Results:** p.N342D caused severe filament-assembly defects with prominent perinuclear aggregates, whereas p.S13F showed mixed phenotypes with frequent perinuclear aggregates, and p.R454W largely preserved filamentous networks. p.N342D and p.S13F induced nuclear deformation with disrupted nesprin-3 and lamin A/C distribution. In p.N342D and p.S13F, desmin aggregates drove focal lamin A/C accumulation, nuclear envelope (NE) rupture, DNA leakage, and increased cGAS activation. *DES* variants significantly shortened APD_20/90_ and reduced CaT amplitude, indicating pro-arrhythmic electrical remodeling. Atrial proteomics revealed a DN–specific signature enriched for cytoskeletal, NE, intermediate filament, and chaperone pathways, consistent with the structural injury observed *in vitro*. GGA prevented desmin aggregation and nuclear morphology changes, and mitigated APD shortening in p.N342D-expressing cardiomyocytes. Human myocardium from *DES* variant carriers showed concordant desmin aggregation and polarized lamin A/C distribution.

**Conclusions:** *DES* variants induce a desmin-dependent atrial cardiomyopathy characterized by cytoskeletal disorganization, disruption of LINC-complex, NE rupture with DNA leakage, and pro-arrhythmic electrophysiological remodeling. These findings provide mechanistic insight into how DN variants promote atrial disease. HSPs induction by GGA partially restores structural and functional integrity, identifying a potential therapeutic approach for desmin-related atrial cardiomyopathy.

**Clinical perspective:** **What is new?**

- Pathogenic *DES* variants induce a previously unrecognized atrial cardiomyopathy characterized by desmin aggregation, and desmin-network (DN) collapse, disruption of the linker of nucleoskeleton and cytoskeleton (LINC) complex, and nuclear envelope rupture with DNA leakage.
- Variants that lead to desmin aggregation (e.g., p.N342D) cause focal lamin A/C polarization, cyclic GMP–AMP synthase (cGAS) activation, and structural injury at the nuclear envelope.
- *DES* variants produce pro-arrhythmic electrical remodeling, including action potential duration shortening and impaired Ca²⁺ handling in HL-1 atrial cardiomyocytes.
- Atrial proteomics from DN variant carriers reveals enrichment of pathways related to cytoskeletal, nuclear envelope, intermediate filament, and chaperone, supporting a desmin-dependent remodeling program.
- The heat-shock protein inducer geranylgeranylacetone (GGA) prevents desmin aggregation, restores nuclear morphology, and mitigates electrical and Ca²⁺ handling remodeling.

**What are the clinical implications?**

- These findings establish DN dysfunction as a distinct cause of atrial cardiomyopathy, providing a mechanistic basis for the association between pathogenic *DES* variants and atrial arrhythmias, including atrial fibrillation.
- Nuclear envelope rupture and cytosolic DNA leakage represent new mechanistic evidence which links cytoskeletal injury and atrial arrhythmogenesis.
- Identifying structural vulnerability in *DES* variant carriers fosters awareness of genetic counseling for atrial disease, enabling early detection and risk stratification.
- The protective effects of GGA suggest that restoring proteostasis may be a therapeutic strategy for desmin-related atrial cardiomyopathy and potentially other genetic atrial diseases.

**Novelty and significance statement:** *Novelty:* This study identifies a desmin-dependent atrial cardiomyopathy driven by cytoskeletal aggregation, LINC-complex disruption, and nuclear envelope rupture with DNA leakage. We show that pathogenic *DES* variants are associated with pro-arrhythmic molecular remodeling and that human atrial proteomics confirm nuclear envelope and cytoskeletal injury as core features. Importantly, the heat-shock protein-inducer GGA rescues structural, molecular, and electrophysiological defects, revealing a modifiable pathway in desmin-mediated atrial disease.

*Significance:* These findings provide the first integrated mechanistic explanation linking DN variants to atrial cardiomyopathy. By uncovering nuclear envelope rupture and cGAS activation as key drivers of atrial cardiomyopathy, this work expands the molecular framework for inherited atrial disease and highlights proteostasis enhancement as a potential therapeutic strategy for patients carrying *DES* and related cytoskeletal variants.

**Graphical abstract:** 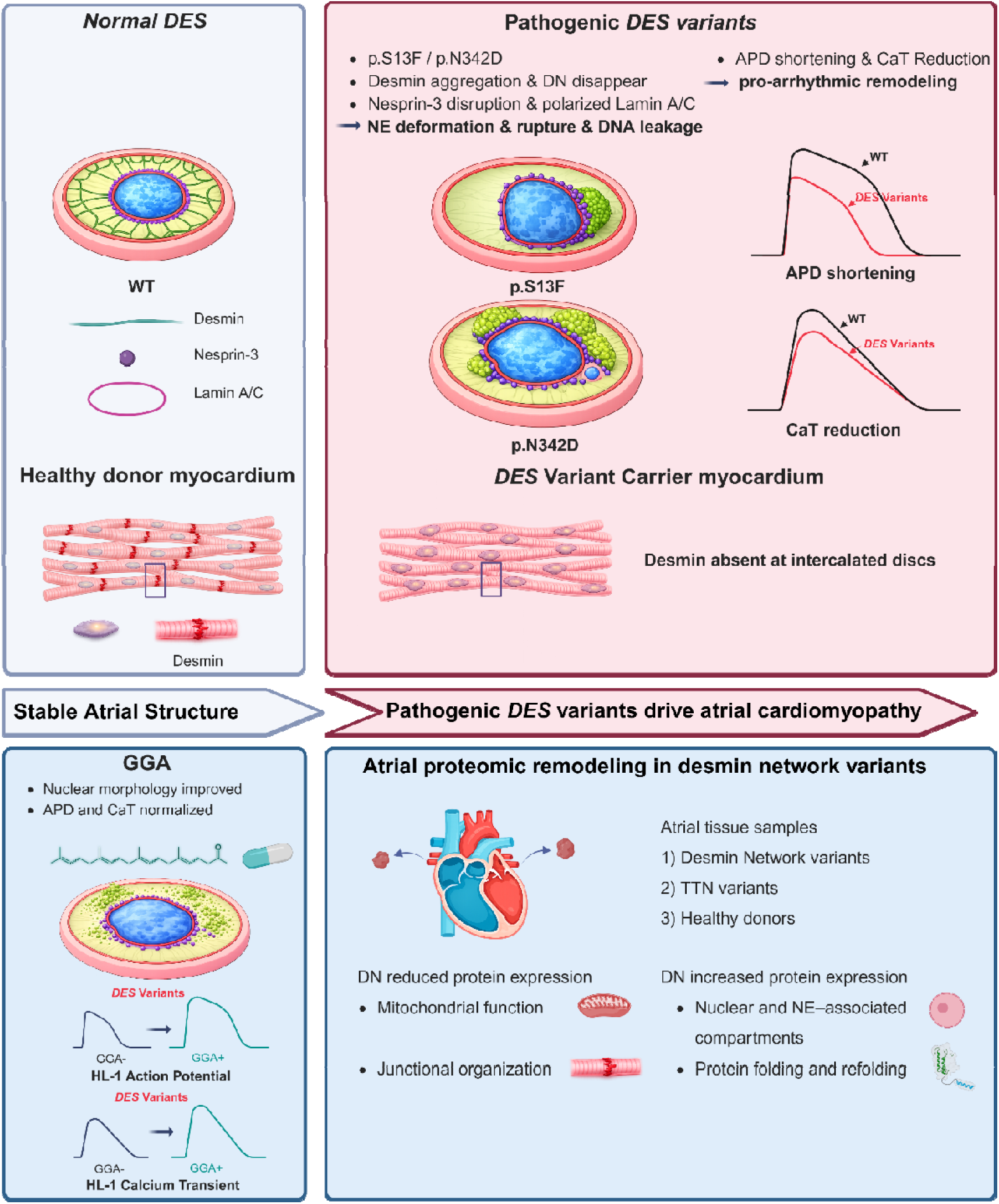

## 1. Introduction

Atrial cardiomyopathy encompasses a spectrum of structural, architectural, contractile, and electrophysiological abnormalities of the atrial myocardium that impair atrial function.^1^ These alterations contribute to the initiation and progression of atrial arrhythmias, including atrial fibrillation (AF).^1,2^ At the cellular level, maintenance of atrial structure and function depends on an integrated network of cytoskeletal and nucleoskeletal systems that ensure mechanical stability and force transmission during cardiac contraction.^3^ This network includes intermediate filaments, desmosomes, microtubules, and the nuclear envelope (NE), which together preserve cardiomyocyte architecture and nuclear integrity. Disruption of these interconnected systems is increasingly recognized as a driver of inherited and acquired atrial disease.^3,4^

Desmin, encoded by the gene *DES*, is the principal intermediate filament in striated muscle and forms a cytoskeletal scaffold linking the sarcomere to mitochondria, intercalated discs (IDs), and the NE^5^ through interactions with plectin and the linker of nucleoskeleton and cytoskeleton (LINC) complex. Through this network, desmin contributes to cytoskeletal organization, nuclear positioning, and mechanotransduction in cardiomyocytes.^6^ Pathogenic variants in *DES* have been found to result in desminopathy,^7^ a myofibrillar myopathy frequently associated with conduction disease, ventricular dysfunction, and arrhythmias.^7,8^ Importantly, several *DES* variants, including p.S13F, p.N342D, and p.R454W, have been identified in individuals with early atrial disease or atrial cardiomyopathy,^8–10^ suggesting that disruption of the desmin-network (DN) may create a pro-arrhythmogenic substrate in atrial myocardium. However, the molecular pathways by which these *DES* variants alter atrial structure and electrophysiological function remain poorly defined.

Although desmin aggregation is a hallmark of desminopathy,^11^ its impact on NE integrity and LINC complex organization in atrial cardiomyocytes remains largely unexplored. NE instability, including lamina deformation, chromatin herniation and rupture, has recently emerged as a key pathogenic mechanism in cardiac and skeletal myopathies.^12,13^ Nuclear rupture permits genomic DNA to leak into the cytosol, where it activates the innate immune DNA sensor by cyclic GMP–AMP synthase (cGAS).^14^ Whether *DES* variants provoke similar nuclear injury in atrial myocardium is unknown. In addition, disruptions in cytoskeletal–nuclear coupling may impair electrophysiological properties, leading to action potential duration (APD) shortening and abnormal calcium handling, features increasingly recognized in atrial cardiomyopathy.^15^ Whether *DES* variants induce comparable electrophysiological remodeling in atrial cardiomyocytes has not been established.

Heat-shock proteins (HSPs) maintain cytoskeletal stability and prevent misfolded protein aggregation.^16^ Pharmacological induction of HSPs using geranylgeranylacetone (GGA) has shown protective effects in experimental models of cardiac stress and cytoskeletal disruption.^17^ Whether HSP induction can mitigate desmin aggregation, NE injury, or electrophysiological remodeling caused by *DES* variants remains unknown.

In this study, we investigated how pathogenic *DES* variants alter atrial cardiomyocyte structure and function. We demonstrate that *DES* variants induce desmin aggregation and disrupt LINC-complex organization, leading to NE rupture with chromatin herniation and DNA leakage accompanied by activation of cGAS. These structural abnormalities are associated with electrophysiological remodeling characterized by altered action potential duration and calcium handling. Human atrial proteomic profiling further revealed a DN specific signature enriched for cytoskeletal, NE, intermediate filament, and chaperone pathways, supporting the relevance of these mechanisms in diseased human atrial myocardium. Importantly, pharmacological induction of HSPs with GGA attenuated desmin aggregation and mitigated structural and electrophysiological abnormalities. Together, these findings identify a desmin-dependent mechanism for atrial cardiomyopathy characterized by cytoskeletal disorganization, disruption of cytoskeletal–nuclear coupling, NE rupture with DNA leakage, and electrophysiological remodeling in atrial cardiomyocytes.

## 2. Materials and Methods

### 2.1 Cell Culture and construction of stable human DES expressing cell lines

HL-1 cardiomyocytes derived from adult mouse atria (Merck, SCC065) were cultured in Claycomb medium (Merck, 51800C) supplemented with 10% FBS (Merck, F9665-500ML), 2 mmol/L L-glutamine (Merck, G7513-100ML), 100□µM norepinephrine (Merck, N5785-1G), and penicillin/streptomycin (Gibco, 15140-122, 1:100).^9^ Human *DES* was cloned into a PiggyBac backbone (pPB533-IRES-neo-WPRE) with an N-terminal FLAG tag. A pmEmerald-Desmin-C-18 plasmid served as the wild-type (WT) template. Missense variants p.S13F and p.R454W were introduced by site-directed mutagenesis (QuikChange XL, Agilent, 200518) using variant-specific oligonucleotides; p.N342D, p.N342E, and p.N342Q were generated by Gibson Assembly (NEB, E2611L). The complete *DES* coding region of each construct was confirmed by Sanger sequencing (Eurofins). Variants were annotated relative to human desmin NM_001927.4. HL-1 cardiomyocyte lines stably expressing *DES-*WT, p.S13F, p.N342D, p.N342E, p.N342Q, or p.R454W were produced by co-transfection of the PiggyBac expression plasmid and transposase helper plasmid using Lipofectamine 3000 (Thermo Fisher Scientific, L3000001). Cardiomyocytes were selected with geneticin (G418, 10131035, 400 µg/mL) from 24 hours post-transfection up to 2 weeks in order to establish stable populations of cardiomyocytes.

To study DNA leakage from nucleus to cytoplasm, HL-1 cardiomyocytes were transiently transfected with pCDH-cGASmut-mNG-Puro using Lipofectamine 3000 (Thermo Fisher Scientific).

### 2.2 GGA treatment of HL-1 cardiomyocytes

HL-1 cardiomyocytes expressing *DES* variants were treated with 10 μM GGA (Chaperone Pharma BV, the Netherlands) or the same volume of DMSO for 48Dh, followed by microscopic analyses and functional measurements.

### 2.3 Molecular modelling

Recently, Eibauer et al. described the molecular structure of the vimentin filaments using cryo-electron microscopy.^18,19^ Due to the high homology of vimentin and desmin especially in the rod- and tail domains, we used this structure (8RVE, https://www.rcsb.org/structure/8RVE) for generation of a desmin model using the SWISS-MODELL server (https://swissmodel.expasy.org/).^20^ Molecular visualization was performed using the PyMOL Molecular Graphics Systems software (Schrödinger LLC, New York, NY, USA).

### 2.4 Protein extraction and Western blot analysis

The BSA protein assay kit (Thermo Fisher Scientific, 23208) was used to estimate the protein content. Subsequently, proteins were separated using sodium dodecyl sulfate–polyacrylamide gel electrophoresis and transferred to PVDF membranes (Merck, IPFL00010). In the Western blot assay, anti-FLAG (Sigma-Aldrich, F7425) and anti-desmin (R&D Systems, AF3844) were used as primary antibodies. Antigen–antibody complexes were detected using the enhanced chemiluminescence detection reagents (Cytiva, 18363513) with anti-goat or anti-rabbit IgG secondary antibodies. Ponceau staining (Thermo Fisher Scientific, A40000278) was used as the endogenous reference.

### 2.5 Confocal microscopy and quantitative image analysis

HL-1 cardiomyocytes were fixed with 4% paraformaldehyde (PFA; Thermo Fisher Scientific, J19943K2) for 10 minutes at room temperature and permeabilized with 0.25% Triton X-100 (Merck, 648463) in PBS for 20 minutes at room temperature. After blocking with 3% bovine serum albumin (BSA) in PBS, the cardiomyocytes were incubated with monoclonal ANTI-FLAG M2 (Merck, F1804-200UG; 1:200), anti-Lamin A/C antibody (Cell Signaling Technology, 4777S; 1:200), anti-desmin (R&D Systems, AF3844; 1:20) or anti-Nesprin-3 antibody (Proteintech, 27132-1-AP; 1:100) overnight at 4°C and were washed with 0.3% BSA/PBS. Next, the cardiomyocytes were incubated with Cy3-conjugated anti–Rabbit-IgG or anti–mouse-IgG secondary antibodies for 1 hour at room temperature. Nuclei were stained with 4′,6-diamidino-2-phenylindole (DAPI; 1:1000; Thermo Fisher Scientific, 62248) for 10 minutes at room temperature, washed with PBS, and imaged using a Nikon confocal microscope (Nikon, Tokyo, Japan).

For immunofluorescence analysis on human cardiac tissue, 10 µm thick paraffin-embedded sections of heart biopsies were deparaffinized by incubation at 70°C for 1 hour and rehydrated with a graded series of ethanol. Antigen retrieval was performed in EDTA (VWR, 33600267) buffer (pH 8.0) using a pressure cooker. Samples were permeabilized with 0.25% Triton X-100 (Merck, 648463) in PBS for 10 minutes at room temperature. After blocking with 3% BSA in PBS for 30 minutes, the sections were incubated with anti-Desmin (AF3844 Bio-Techne, 10 µg/mL), and anti-Lamin A/C antibody (Cell Signaling Technology, 4777S 1:200) and then with Alexa Fluor-conjugated secondary antibodies (Life Technologies). Nuclei were stained with DAPI (1:1000 in PBS, 10 min), washed three times in PBS, and mounted in Mowiol on glass slides. Imaging parameters matched those used for HL-1 cardiomyocytes. Frozen left ventricular cryosections (10 μm) were air-dried, fixed in 4% PFA for 10 minutes at room temperature, washed in PBS, permeabilized with 0.25% Triton X-100 for 10 minutes, and blocked with 3% BSA for 30 minutes. Primary antibody incubation was performed overnight at 4°C, and subsequent secondary antibody incubation, nuclear counterstaining, mounting, and imaging were carried out as described above for FFPE sections.

Nuclear morphology was quantified using Fiji. DAPI-stained nuclei were acquired as 16-bit images under identical settings (×200), converted to 8-bit, and smoothed using a Gaussian blur (σ = 2). Automated thresholding (Default dark) was applied to generate binary masks, followed by hole filling. Nuclei were segmented using Analyze Particles with exclusion of edge objects and a minimum size threshold of ≥20 pixels. Nuclear area, perimeter, and circularity (4π × area/perimeter²) were quantified.

To assess lamin A/C distribution at the nuclear envelope, a line was drawn across the nucleus from the nuclear rim opposite the desmin aggregate through the nuclear center toward the region adjacent to the desmin aggregate. Fluorescence intensity along the line was quantified using the “Plot Profile” function in FIJI. Intensity values were exported to Microsoft Excel, where the two fluorescence maxima corresponding to the nuclear rims were identified as peaks. The distance between these peaks was normalized to generate a standardized x-axis for all nuclei analyzed. Fluorescence intensity values were normalized to the maximal lamin A/C signal at the nuclear rim opposite the desmin aggregate site to enable comparison of lamin A/C distribution between cardiomyocytes.

### 2.6 HL-1 cardiomyocyte calcium and action potential measurements

To load cardiomyocytes with the Ca^2+^ indicator Fluo-4 AM (Thermo Fisher, F14201), Fluo-4 AM was dissolved in DMSO to a final concentration of 1 mM. The Fluo-4 AM solution was further diluted in 1 mL Opti-MEM (Thermo Fisher, 31985070) to a final concentration of 2 µmol/L. In the same solution, FluoVolt™ and PowerLoad™ (Thermo Fisher, F10488) were diluted at 1:1000 and 1:100 respectively. After 30 min incubation in the dark at 37°C, the HL-1 cardiomyocytes were washed three times with Opti-MEM for 5 minutes each. Dye de-esterification was allowed to occur during a subsequent 15-minute incubation in fresh Opti-MEM.

HL-1 cardiomyocytes were loaded into the CytoCypher MultiCell system and were electrically paced via a MyoPacer (IonOptix) at 1 Hz at 40 V for 20 ms and fluorescent signals were recorded using IonWizard data acquisition software (IonOptix). Calcium transient (CaT) parameters were analyzed from normalized fluorescence signals (F/F□). APD at 20% and 90% repolarization were determined. For each cardiomyocyte, 10 consecutive steady-state waveforms were averaged to obtain a single value per cell. These cell-level values were then averaged per independent experiment.

### 2.7 Clinical description of cardiac tissue samples and ethics

Human myocardial tissue from heterozygous *DES* variant carriers was obtained from two independent centers.

Explanted left ventricular (LV) myocardial tissue from a carrier of the *DES* p.R454W variant was obtained at the time of cardiac transplantation at the Heart and Diabetes Center North Rhine-Westphalia, Ruhr-University Bochum, Bad Oeynhausen, Germany. The collection and use of these samples for research purposes were approved by the Ethics Committee of the Ruhr-University Bochum (Bad Oeynhausen, Germany; approval number AZ 2024-1256). Written informed consent was obtained from the patient.

For *DES* p.S13F and p.P419R variant carriers, myocardial tissue was obtained through the TransplantLines Biobank and Cohort Study at the University Medical Center Groningen, University of Groningen, Groningen, The Netherlands (ClinicalTrials.gov identifier: NCT03272841). The TransplantLines study protocol was approved by the Medical Ethics Review Board of the University Medical Center Groningen, The Netherlands. All participants provided written informed consent prior to inclusion in the study.

TransplantLines is an ongoing, prospective study aiming to improve understanding of disease-related and ageing-related outcomes and health problems in solid organ transplant recipients and donors. From June 2015, all (potential) solid organ transplantation patients and (potential) living organ donors (aged ≥18 years) at UMCG were invited to participate. All participants provided written informed consent at enrollment. A detailed description of the study design, inclusion and exclusion criteria has been published previously.^21^ The study protocol was approved by the local Institutional Review Board (METc 2014/077), adheres to the UMCG Biobank Regulation, and complies with the WMA Declaration of Helsinki and the Declaration of Istanbul. Myocardial samples from UMCG were formalin-fixed, paraffin-embedded (FFPE) and sectioned at 10 µm. Frozen LV cryosections from the p.R454W carrier were processed separately according to standard immunofluorescence protocols.

### 2.8 Proteomics of human atrial tissue samples

Human right and left atrial tissue samples from patients with end stage heart failure carrying variants in DN associated genes (*DES, DSG, PKP2, CRYAB, DSP*; n=7), *TTN* variants (n=5), and from non-diseased rejected donor hearts (control) were lysed and digested with trypsin in a Barocycler 2320XT (Pressure Biosciences). Resulting peptides were purified and desalted on C18 tips and spiked with a peptide retention time calibration standard (iRT, Biognosys). For quantitative proteome profiling, 400 ng protein equivalent of digested peptides were analyzed by data-independent acquisition mass spectrometry (DIA-MS) on a hybrid ion mobility/quadrupole/time-of-flight mass spectrometer (timsTOF Pro 2, Bruker) using a 100 min gradient and a custom-made 20×2 variable window diaPASEF acquisition method.^22,23^ Two to three mass spectrometry replicates per sample were acquired.

Raw data were processed in Spectronaut v19.2 software.^23^ Protein detection was performed against an in-house deep spectral library generated from human cardiac tissues using the Pulsar search engine and the UniProtKB human reference proteome v8.2023, with default parameters and an additional 55-protein in-house contaminant database. Precursor, peptide, and proteins false discovery rates were controlled at 1% using a concatenated forward-and-reverse decoy database approach. DIA quantification used up to six fragment ions per peptide and up to ten peptides per protein, with dynamic retention time alignment, dynamic mass recalibration to align feature matching. Quantitative values were quartile normalized, and global imputation using default settings applied for the final results table.

### 2.9 Statistical analyses

Statistical analyses were performed using GraphPad Prism 10.6.0. Data are presented as mean ± SD, gray dots represent individual nuclei or cardiomyocytes, and colored dots indicate the mean of each independent experiment. *n* denotes independent biological replicates (typically n=3), each containing multiple technical measurements. Normality was assessed by Shapiro–Wilk testing. Comparisons among *DES* variants (WT, p.S13F, p.N342D, p.R454W) were made by using one-way ANOVA with Dunnett’s post-hoc test versus WT. Lamin A/C intensity between aggregate-facing and non-aggregate nuclear sides was compared using a paired t test. GGA intervention effects were analyzed by two-way repeated-measures ANOVA with Tukey’s multiple-comparisons test for prespecified contrasts. All tests were two-sided, and *p* < 0.05 was considered statistically significant.

For DIA-MS proteomics, statistical analyses and visualizations (PCA, volcano plots) were performed in Origin-Pro 8.5G and eVITTA v1.3.1. Differentially abundant proteins (DAPs) were defined as FDR-adjusted *p* < 0.05 with fold changes ≥2.0 or ≤0.5. Functional enrichment analysis of DAPs was conducted using Metascape (https://metascape.org) with default parameters, with annotations restricted to Gene Ontology Molecular Function (GO-MF), Cellular Component (GO-CC), Biological Process (GO-BP), and KEGG pathway analyses.

## 3. Results

### 3.1 Pathogenic human DES variants promote desmin aggregate formation in cardiomyocytes

To investigate how pathogenic *DES* variants contribute to atrial cardiomyopathy, we expressed three clinically established *DES* missense variants, p.S13F (N-terminal head domain), p.N342D (rod domain, segment 2B), and p.R454W (C-terminal tail domain), in HL-1 atrial cardiomyocytes (Figure 1A). Also, a molecular model was generated of the desmin antiparallel tetramer based on the recently resolved cryo-electron microscopy structure of the highly homologous intermediate filament protein vimentin (Figure 1B). These variants were selected because they span the major functional domains of desmin and have been associated with early-onset cardiac conduction disease and atrial and ventricular arrhythmias, including AF, in multiple independent clinical cohorts.^8^ Prior clinical series in Dutch founder cohorts established high cardiac penetrance for p.S13F and p.N342D with early atrial arrhythmia onset,^9,24^ whereas p.R454W has been repeatedly associated with severe cardiomyopathy accompanied by arrhythmias.^10^

**Figure 1.**
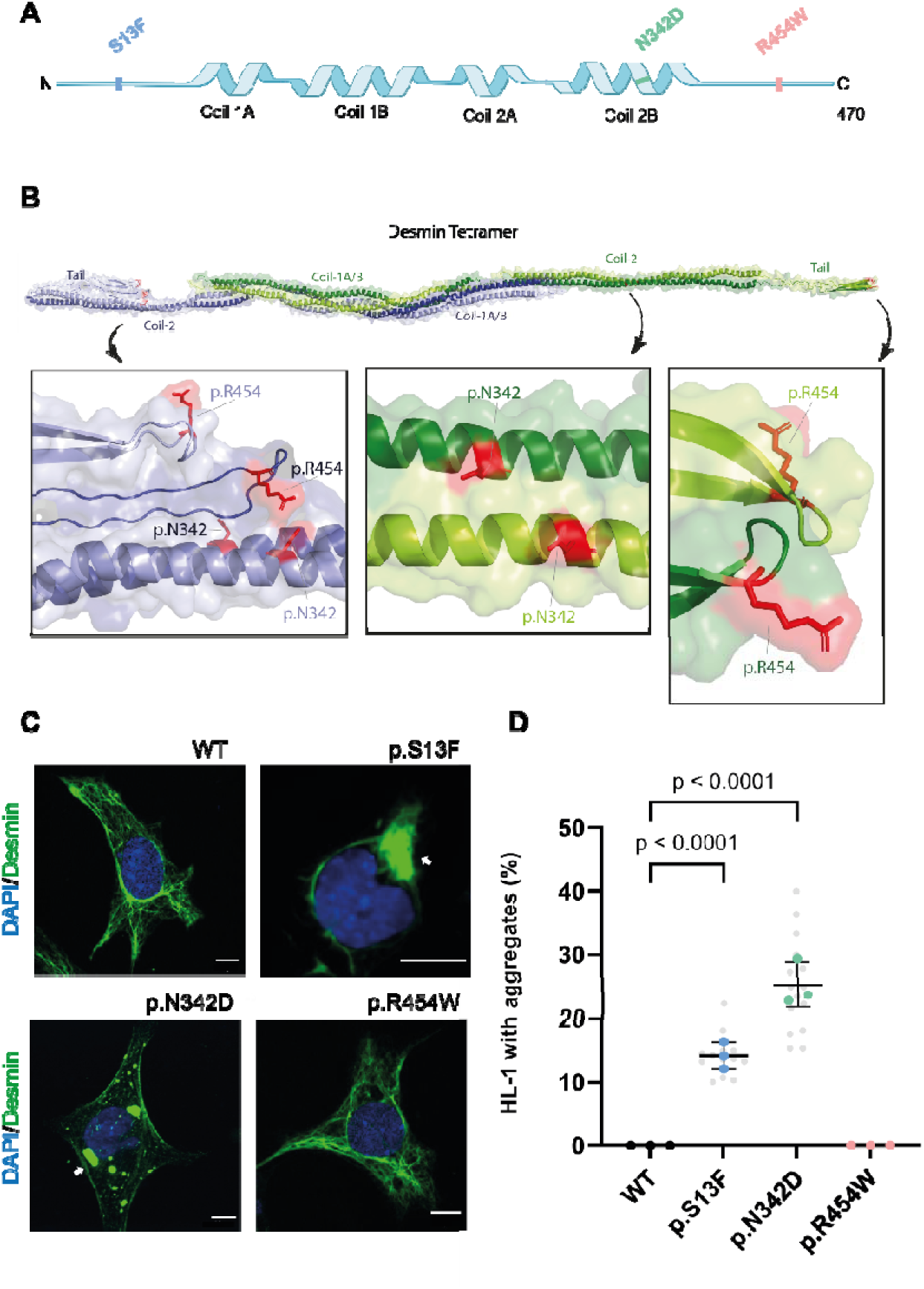
Schematic domain structure of desmin and desmin aggregation in HL-1 atrial cardiomyocytes. **A,** Location of the variants are marked: p.S13F in the head (blue), p.N342D in the rod domain (green), and p.R454W in the tail (pink). **B,** A molecular model of the antiparallel non-symmetric desmin tetramer formed by two coiled-coil dimers (green and blue) is shown. Asparagine-342 (N342) and arginine-454 (R454W) residues are shown as sticks (red) and are labeled in the color of the corresponding helices. N342 is localized in the coil-2 region of the rod-domain, whereas R454 is localized at a loop within the tail domain. Due to back folding of the tail-domains these two amino acids are in close proximity in the antiparallel dimer. **C**, Representative confocal images of HL-1 cardiomyocytes stably expressing *DES*-WT, p.S13F, p.N342D, or p.R454W. Desmin (green) shows an extended cytoplasmic filament network in WT and p.R454W, whereas p.S13F and p.N342D frequently display compact perinuclear aggregates (arrows). Nuclei are counterstained with DAPI (blue). Scale bars, 10 μm. **D**, Percentage of desmin aggregates in HL-1 cardiomyocytes expressing *DES*-WT, p.S13F, p.N342D, or p.R454W. Each gray dot represents one image field; colored dots (3 per group) represent the mean of each independent experiment (n=3); bars show mean±SD. In total, *DES*-WT n=105, p.S13F n=95, p.N342D n=101, and p.R454W n=99 cardiomyocytes were analyzed (5 fields per construct per cell line, ≥15 cardiomyocytes per field). Statistics: one-way ANOVA with Dunnett’s multiple comparisons vs WT.

Because HL-1 cardiomyocytes express endogenous desmin, desmin organization analyses were performed using anti-FLAG immunostaining, thereby selectively visualizing expression of exogenous human desmin. Parental HL-1 cardiomyocytes showed no FLAG signal, confirming antibody specificity (Figure S1A). Western blot analysis further validated expression of exogenous desmin constructs using anti-FLAG antibody, while total desmin immunoblotting detected both endogenous and exogenous desmin (Figure S1B). These data confirm successful expression of human DES variants in HL-1 cardiomyocytes.

Next, desmin aggregate formation was assessed in HL-1 atrial cardiomyocytes. WT cardiomyocytes displayed a filamentous cytoplasmic DN (Figure 1C). A comparable filamentous organization of desmin was observed in cardiomyocytes expressing p.R454W, with no apparent disruption or loss of filament integrity. In contrast, expression of p.S13F and p.N342D resulted in the formation of compact, predominantly perinuclear desmin aggregates (Figure 1C). Quantitative analysis demonstrated desmin aggregation in 25.2% of p.N342D-expressing cardiomyocytes and in 14.2% of p.S13F-expressing cardiomyocytes (Figure 1D), indicating impaired filament assembly associated with these variants. In contrast to human ventricular tissue samples of p.R454W carriers,^25^ stable expression of p.R454W in HL-1 cardiomyocytes did not induce aggregation, suggesting that additional factors may contribute to filament instability or aggregation in human cardiac muscle.

As residue N342 is located within the highly conserved rod domain (coil 2B) and appeared particularly aggregation-prone, we next investigated whether the physicochemical properties of the side chain at this position determine filament behavior. Based on prior reports that rod-domain *DES* variants disrupt filament assembly and promote aggregation, we hypothesized that introducing an acidic residue at position 342 would alter desmin assembly and aggregation propensity.^26^ To directly test this concept in an atrial cardiomyocyte context, we generated additional HL-1 cardiomyocytes stably expressing p.N342Q (amide), p.N342D (acidic, short side chain), and p.N342E (acidic, longer side chain), and analyzed desmin organization by confocal microscopy. Contrary to our initial hypothesis, p.N342Q induced robust perinuclear aggregation comparable to p.N342D, whereas p.N342E largely preserved a filamentous network (Figure S2). These findings indicate that aggregation propensity at residue 342 is not dictated solely by amide versus acidic side-chain chemistry, but is likely influenced by more subtle structural or steric effects within the coiled-coil domain.

### 3.2. Effect of DES variants on nuclear morphology

The impact of pathogenic *DES* variants on nuclear morphology was examined in HL-1 cardiomyocytes by confocal microscopy, followed by quantitative analysis of nuclear morphology parameters (Figure 2A). Compared to WT, expression of p.S13F and p.N342D was associated with a significant reduction in nuclear circularity (Figure 2B), whereas nuclear perimeter and area were comparable between groups (Figure 2C, D). These alterations indicate a disruption of nuclear shape integrity and suggest that the p.S13F and p.N342D variants affect nuclear–cytoskeletal coupling, contributing to structural abnormalities at the cellular level.

**Figure 2.**
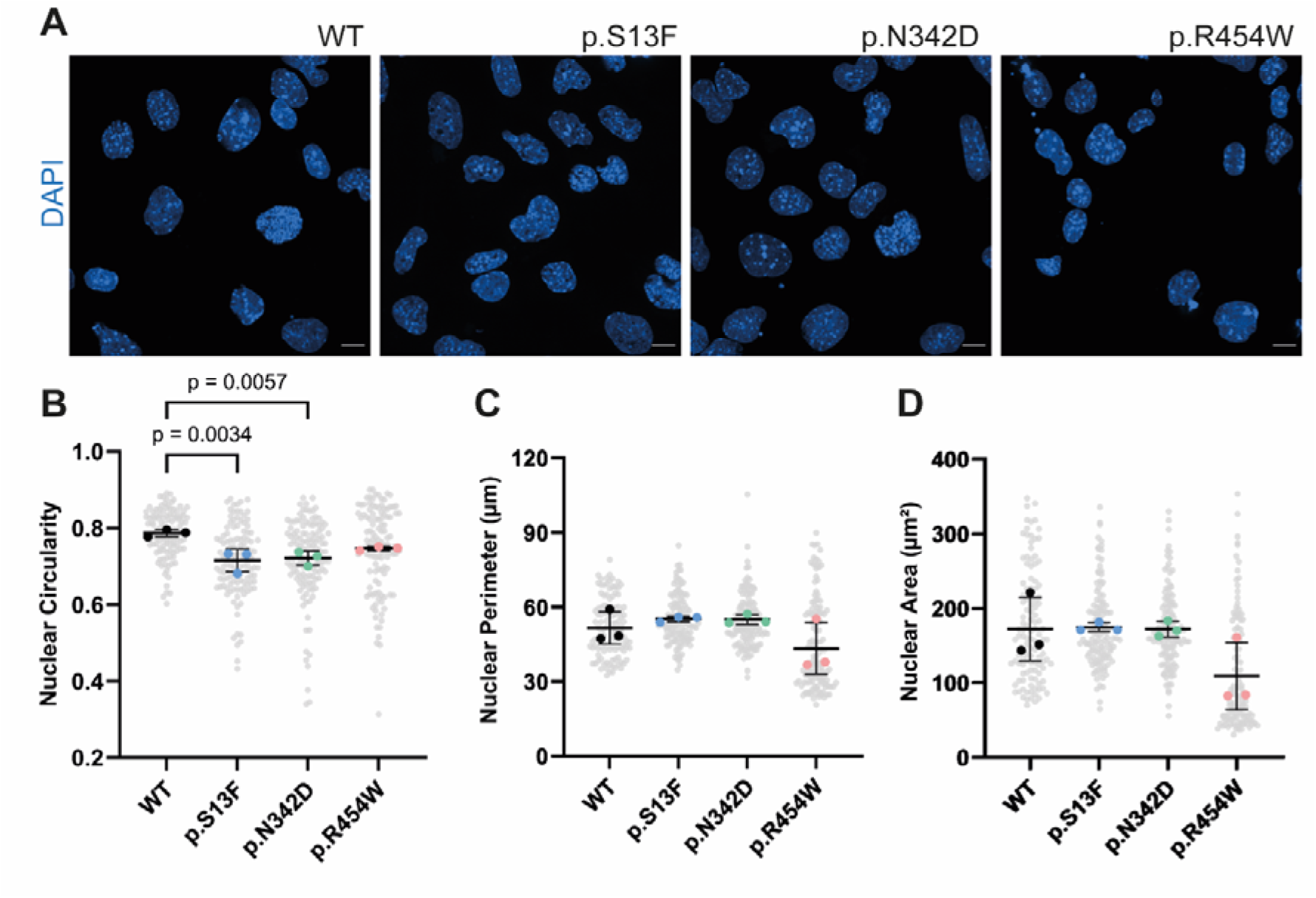
Desmin variants alter nuclear morphology in HL-1 cardiomyocytes. **A**, Representative DAPI-stained nuclei from HL-1 cardiomyocytes expressing *DES*-WT, p.S13F, p.N342D, or p.R454W. Scale bars, 10 μm. **B–D**, Quantification of nuclear circularity (B), perimeter (C), and area (D) from automated segmentation. Gray dots represent individual nuclei; colored dots (3 per group) represent the mean of each independent experiment (n=3); bars show mean±SD across experiments. In total, WT n=114, p.S13F n=121, p.N342D n=125, and p.R454W n=118 nuclei were analyzed. Statistics: one-way ANOVA with Dunnett’s multiple comparisons vs WT.

### 3.3 DES variants remodel the nuclear envelope and disrupt the desmin–nesprin-3 network

Lamin A/C distribution was analyzed in HL-1 cardiomyocytes expressing *DES*-WT, p.S13F, p.N342D, or p.R454W to determine whether *DES* variant–associated aggregates remodel the NE by altering lamin A/C organization. In *DES*-WT cardiomyocytes, lamin A/C formed a smooth, continuous rim with largely homogeneous intensity around the NE (Figure 3A). A similar uniform lamin A/C distribution was observed in p.R454W-expressing cardiomyocytes (Figure 3A). In contrast, in p.S13F and p.N342D expressing cardiomyocytes, lamin A/C intensity was frequently enriched at the NE regions adjacent to perinuclear desmin aggregates, as visualized by lamin A/C heat maps (Figure 3A). This asymmetry was quantified by drawing a directional line scan across the nuclear rim from the non-aggregate side toward the aggregate-facing region (Figure 3B, E). Averaged normalized line-scan profiles showed a clear lamin A/C intensity peak at the aggregate-facing segment in both p.S13F and p.N342D (Figure 3C, F). For quantitative comparison, mean lamin A/C fluorescence was calculated for the non-aggregate rim and the aggregate-facing segment within each nucleus. Lamin A/C intensity was significantly higher at the aggregate-facing region in p.S13F (Figure 3D, *p* < 0.01) and in p.N342D (Figure 3G, *p < 0.05*). For each construct, ten nuclei per experiment were analyzed across three independent experiments (n=30 nuclei total). Together, these results demonstrate that *DES* variants associated with perinuclear aggregation (p.S13F and p.N342D) induce polarized redistribution of lamin A/C at the NE, whereas *DES*-WT and p.R454W maintain a more uniform lamina organization.

**Figure 3.**
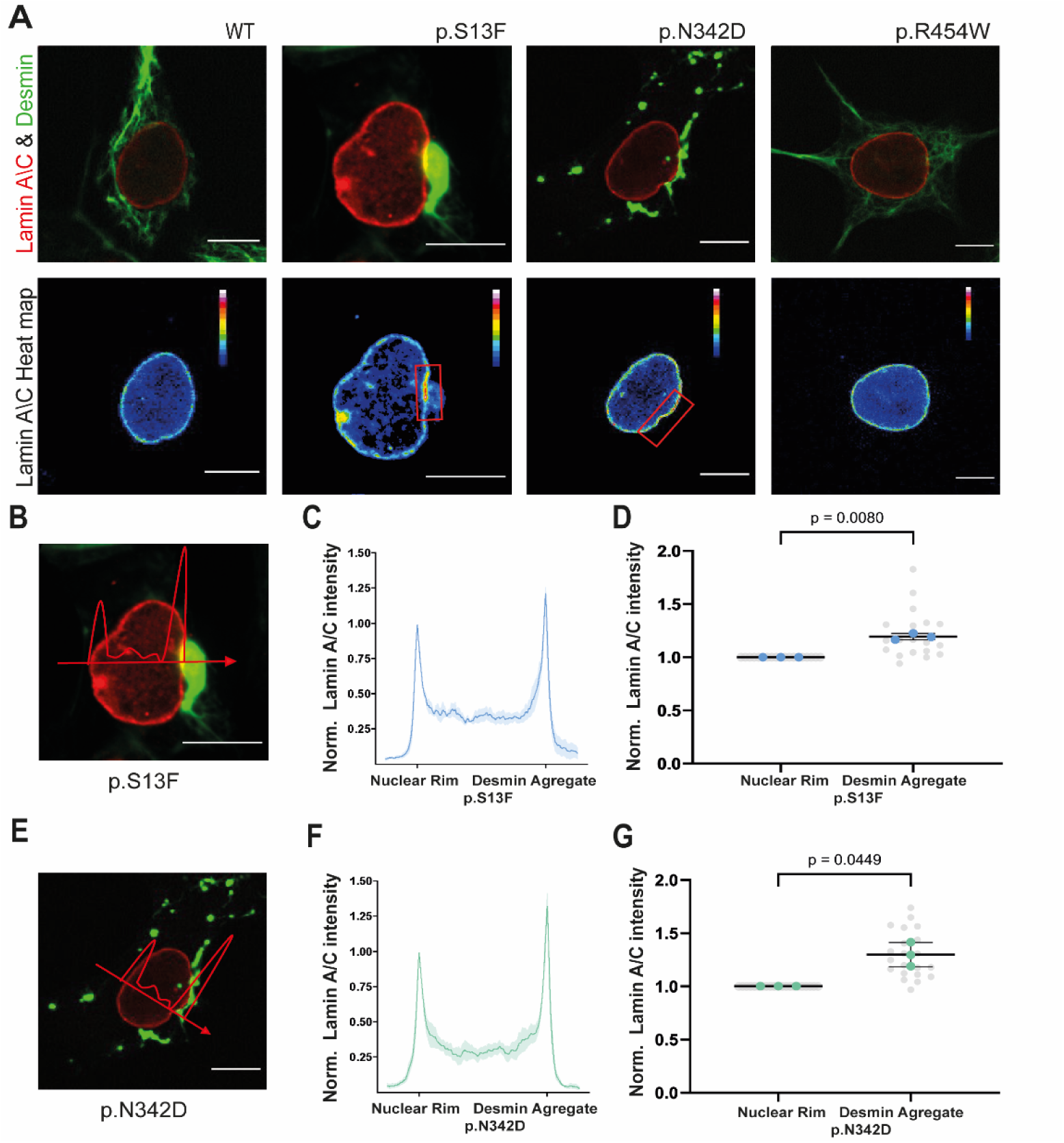
Perinuclear desmin aggregation is associated with polarized lamin A/C distribution. **A**, Representative images of *DES*-WT, p.S13F, p.N342D, and p.R454W HL-1 cardiomyocytes stained for lamin A/C (red) and desmin (green), with corresponding lamin A/C heat maps. WT and p.R454W show a uniform lamin A/C rim, whereas p.S13F and p.N342D display focal lamin A/C enrichment at nuclear regions adjacent to perinuclear desmin aggregates (red boxes). Scale bars, 10 μm. **B**, **E**, Zoomed nuclei from p.S13F (B) and p.N342D (E) with line-scan paths (red) drawn from the non-aggregate nuclear rim toward the desmin aggregate. **C**, **F**, Averaged, normalized lamin A/C fluorescence profiles along the line scans in B and E, showing peaks at the aggregate-facing regions. Shading = SEM. **D**, **G**, Normalized lamin A/C intensity at the non-aggregate rim vs aggregate-facing segment for p.S13F (D) and p.N342D (G). For p.S13F, n=19 nuclei were analyzed; for p.N342D, n=21 nuclei were analyzed, pooled from 3 independent experiments. Gray dots represent individual nuclei; colored dots represent experiment means; bars show mean±SD. Statistics: paired *t* test for rim vs desmin-aggregate–facing segments.

Because desmin interacts with the LINC complex through plectin–nesprin-3 connections,^27^ we next examined whether desmin aggregation also affects nesprin-3 organization. In *DES*-WT cardiomyocytes, nesprin-3 formed a smooth, continuous rim along the NE, consistent with intact LINC complex organization (Figure S3). A similar nesprin-3 pattern was observed in most p.R454W-expressing cardiomyocytes, which largely preserved a filamentous desmin network. In contrast, cardiomyocytes expressing p.S13F and p.N342D frequently exhibited clustered, irregular, and partially fragmented nesprin-3 staining, with gaps in the nuclear rim and focal accumulations adjacent to perinuclear desmin aggregates (Figure S3). Although some p.S13F cardiomyocytes retained a near-normal rim, a substantial fraction displayed nesprin-3 disorganization similar to that observed in p.N342D, particularly when aggregates were located near the NE. These findings indicate that aggregation-prone *DES* variants destabilize the perinuclear desmin scaffold and disrupt the nesprin-3–containing LINC complex.

### 3.4 DES variants induce nuclear envelope rupture

Given the pronounced nuclear deformation induced by specific *DES* variants, NE rupture was assessed as a potential underlying mechanism. NE rupture is known to activate innate immune signaling pathways, including those mediated by the cytosolic DNA sensor cGAS.^14^ Detection of cGAS accumulation at discrete cytoplasmic or perinuclear sites is commonly interpreted as evidence of nuclear DNA leakage into the cytoplasm following NE disruption. Accordingly, cardiomyocytes expressing mNG-cGAS and *DES*-WT, p.S13F, p.N342D, and p.R454W were stained for lamin A/C. In *DES*-WT and p.R454W cardiomyocytes (Figure 4A–D, A3–D3), lamin A/C formed a smooth and continuous nuclear rim, while mNG-labeled cGAS was largely diffuse. In contrast, cardiomyocytes expressing p.S13F or p.N342D frequently exhibited discrete cGAS-positive foci, often localized near the NE and adjacent to sites of nuclear blebbing (arrows in Figure 4C1–C2). Quantitative analysis of cGAS-foci–positive cardiomyocytes demonstrated a significant increase in both p.S13F- and p.N342D-expressing cardiomyocytes compared with *DES*-WT, whereas p.R454W did not differ from WT (Figure 4E). These findings indicate that *DES* variants associated with pronounced NE disruption, particularly p.S13F and p.N342D, are linked to increased cGAS accumulation consistent with NE rupture, while p.R454W does not measurably induce this response under the experimental conditions used.

**Figure 4.**
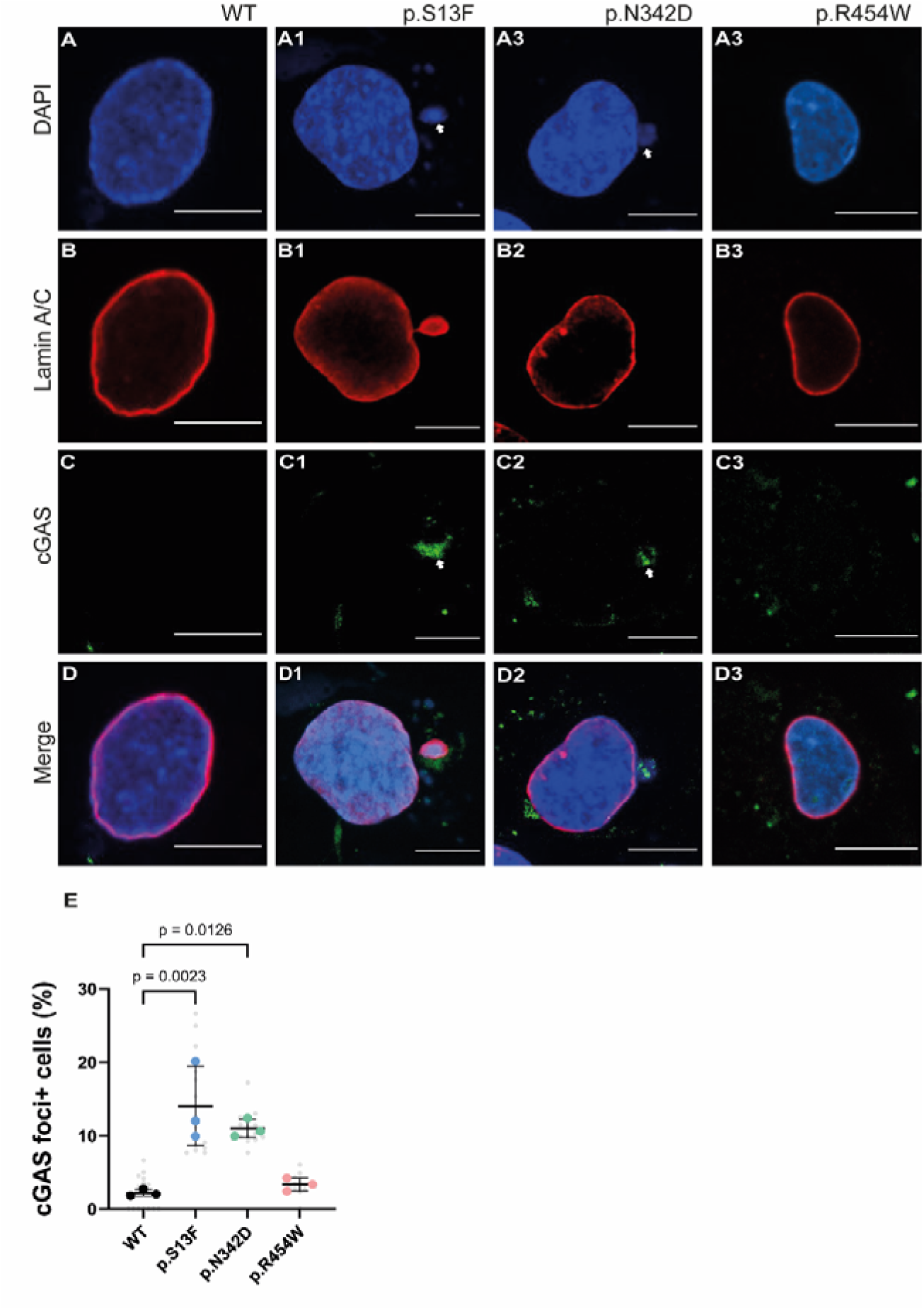
DES aggregation-prone variants increase cyclic GMP–AMP synthase (cGAS) foci consistent with nuclear envelope rupture. **A–D**, Representative confocal images of HL-1 cardiomyocytes expressing *DES*-WT, p.S13F, p.N342D, or p.R454W. **A–A3**, DAPI (blue). **B–B3**, Lamin A/C (red). **C–C3**, cGAS (green). **D–D3**, Merged channels. Arrows indicate discrete cGAS puncta near the NE. Scale bars, 10 μm. **E**, Percentage of cGAS-foci–positive cardiomyocytes (≥1 cGAS punctum per nucleus). For each image, cGAS-positive cardiomyocytes were divided by total DAPI-stained cardiomyocytes to obtain an image-level percentage; image-level values were averaged within each experiment to yield one value per construct. Gray dots represent images; colored dots represent experiment means (n=3); bars show mean±SD. Total cardiomyocytes analyzed: WT n=376, p.S13F n=287, p.N342D n=345, p.R454W n=369. Statistics: one-way ANOVA with Dunnett’s multiple comparisons vs WT.

### 3.5 DES variants alter action potential and calcium handling properties in HL-1 cardiomyocytes

To evaluate the impact of *DES* variant expression on functional outcomes in cardiomyocytes, we analyzed the action potentials and CaT in HL-1 cardiomyocytes expressing either *DES* WT or p.S13F, p.N342D and p.R454W. Relative to WT, p.N342D exhibited shorter APD□□ (Figure 5B) and APD□□ (Figure 5C), with p.S13F and R454W shorter APD□□. The normalized Δvoltage fluorescence amplitude was decreased in *DES* variants p.N342D (Figure 5F). Consistent with these voltage changes, CaT were blunted (Figure 5D). In addition, CaT amplitude (F1/F0) was significantly reduced in p.S13F and p.N342D (Figure 5E). These findings suggest that all three studied *DES* variants disrupt electrophysiological properties and p.S13F, p.N342D variants impair calcium-handling in atrial cardiomyocytes, as such creating an atrial cardiomyopathy phenotype.

**Figure 5.**
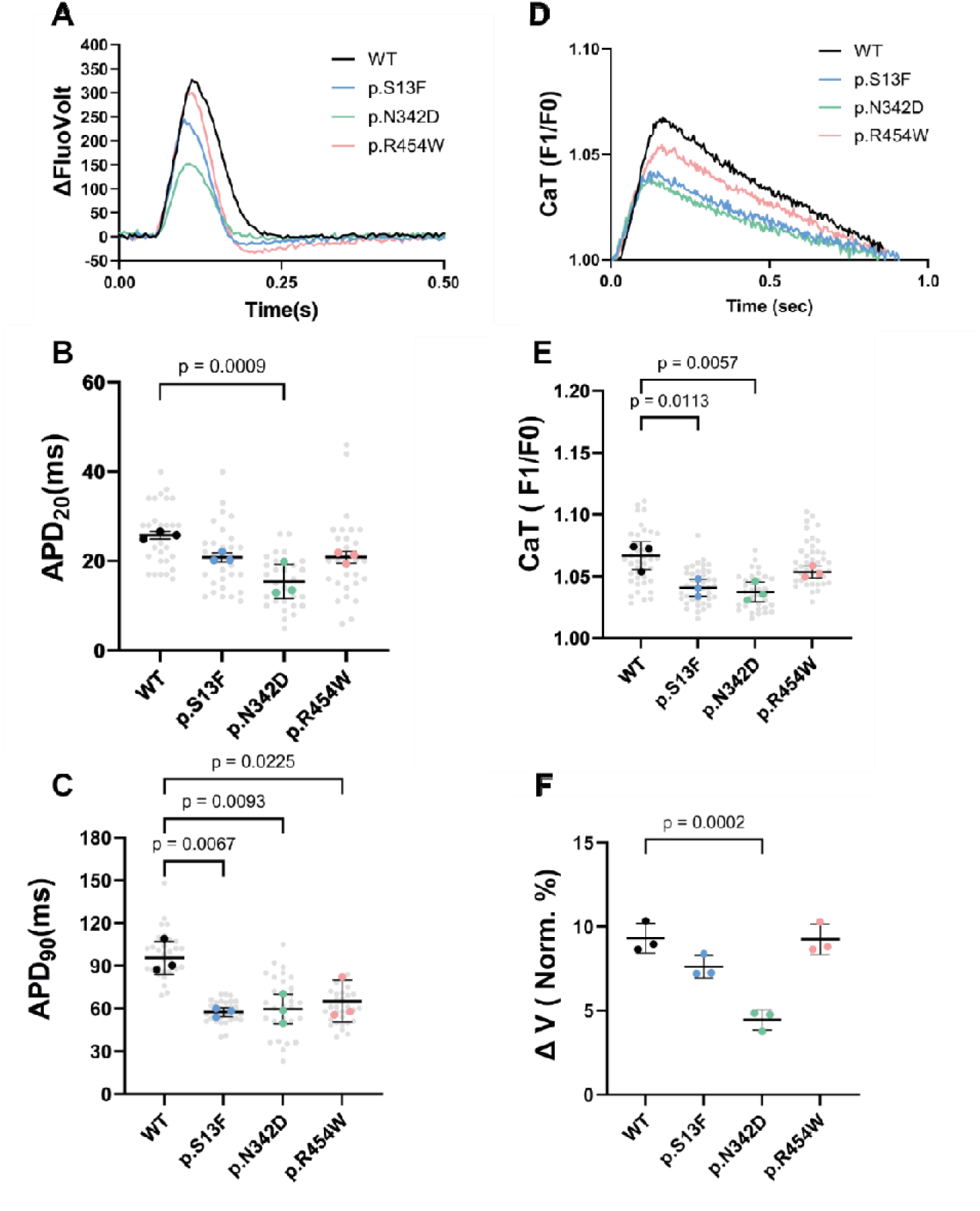
Desmin variants modulate action potentials and Ca²⁺ handling in HL-1 cardiomyocytes. **A**, Representative action-potential traces (FluoVolt ΔV) from *DES*-WT, p.S13F, p.N342D, or p.R454W paced at 1 Hz. **B**-**C**, Action-potential duration at 20% and 90% repolarization (APDLJLJ, APDLJLJ). **D**, Representative CaT traces (F1/F0) from the same constructs. **E**, Peak calcium transients (CaT) amplitude (F1/F0). **F**, Normalized Δ-voltage fluorescence amplitude (% of baseline). Gray dots represent individual cardiomyocytes; colored dots (3 per group) represent the meaning of each independent experiment (n=3); bars show mean±SD. Statistics: one-way ANOVA with Dunnett’s multiple comparisons vs WT.

### 3.6 GGA reduces desmin aggregation and preserves electrophysiology in DES variant HL-1 cardiomyocytes

GGA is a compound that increases expression levels of HSPs^7^ that are known to guide protein folding and refolding, stabilize misfolded proteins, prevent protein aggregation, and facilitate degradation of damaged proteins.^15^ Because it is unknown whether GGA can rescue desmin aggregation in cardiomyocytes. We therefore treated HL-1 cardiomyocytes expressing *DES* variants with 10□μM GGA or vehicle (DMSO) for 48 h followed by measurement of aggregate formation. We observed that GGA significantly reduced desmin aggregation in the p.S13F and p.N342D variants compared to the DMSO control (Figure 6A). Additionally, GGA altered nuclear circularity in the p.N342D variant, while no significant effect was observed in p.S13F (Figure 6B).

**Figure 6.**
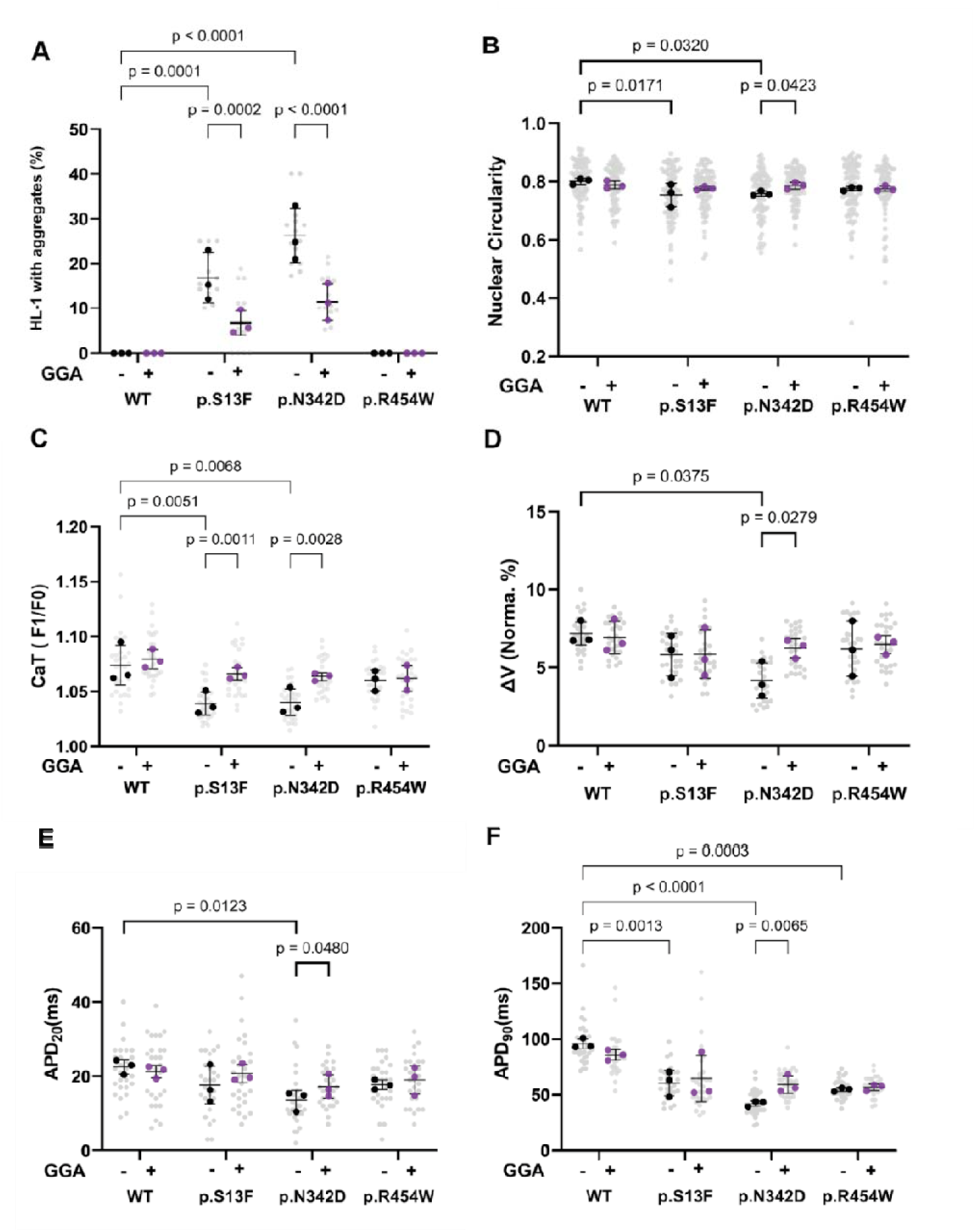
Geranylgeranylacetone reduces aggregate formation and restores electrophysiological and calcium handling properties in DES variant cardiomyocytes. HL-1 cardiomyocytes expressing *DES*-WT, p.S13F, p.N342D, or p.R454W were treated with 10 μM GGA or vehicle (DMSO) for 48 h. **A**, Percentage of cardiomyocytes with desmin aggregates. **B**, Nuclear circularity. **C**, CaT amplitude (F1/F0). **D**, Normalized Δ-voltage fluorescence amplitude. **E**-**F**, Action-potential duration at 20% and 90% repolarization (APDLJLJ, APDLJLJ). Gray dots represent individual cardiomyocytes; colored dots (3 per group) represent the mean of each independent experiment; bars show mean±SD. Two-way ANOVA with Šídák’s multiple comparisons test for the indicated comparisons (within-genotype +GGA vs −GGA; selected comparisons vs WT).

To study whether GGA treatment protects against *DES* variant-induced functional changes, HL-1 cardiomyocytes expressing *DES* variants were treated with GGA or the same volume of DMSO for 48□h. GGA prevented the reduction in CaT amplitude in the p.S13F and p.N342D variants (Figure 6C). GGA also preserved normalized Δ-voltage fluorescence intensity in p.N342D (Figure 6D), indicating an effect on membrane voltage signaling. Additionally, GGA mitigated early repolarization defects (APD_20_) in p.N342D (Figure 6E), and protected against APD_90_ shortening in the p.N342D variant (Figure 6F).

These findings suggest that GGA exerts a protective effect against electrophysiological dysfunction associated with *DES* variants.

### 3.7 Morphological confirmation of desmin and nuclear envelope alterations in human myocardium of DES variant carriers

Myocardial tissue from heterozygous carriers of *DES* variants p.S13F, p.P419R, and p.R454W was analyzed to determine whether the structural abnormalities observed in HL-1 cardiomyocytes are also present in human cardiac tissue.

Immunofluorescence analysis demonstrated variant-dependent disruption of desmin organization. In the p.S13F carrier, focal desmin aggregates were observed, while many cardiomyocytes retained a partially filamentous network (Figure 7A1). In contrast, myocardium from the p.P419R carrier exhibited diffuse, high-intensity desmin aggregation with marked loss of filament organization (Figure 7A2). The p.R454W sample showed desmin aggregates with partial preservation of filament structure (Figure 7A3).

**Figure 7.**
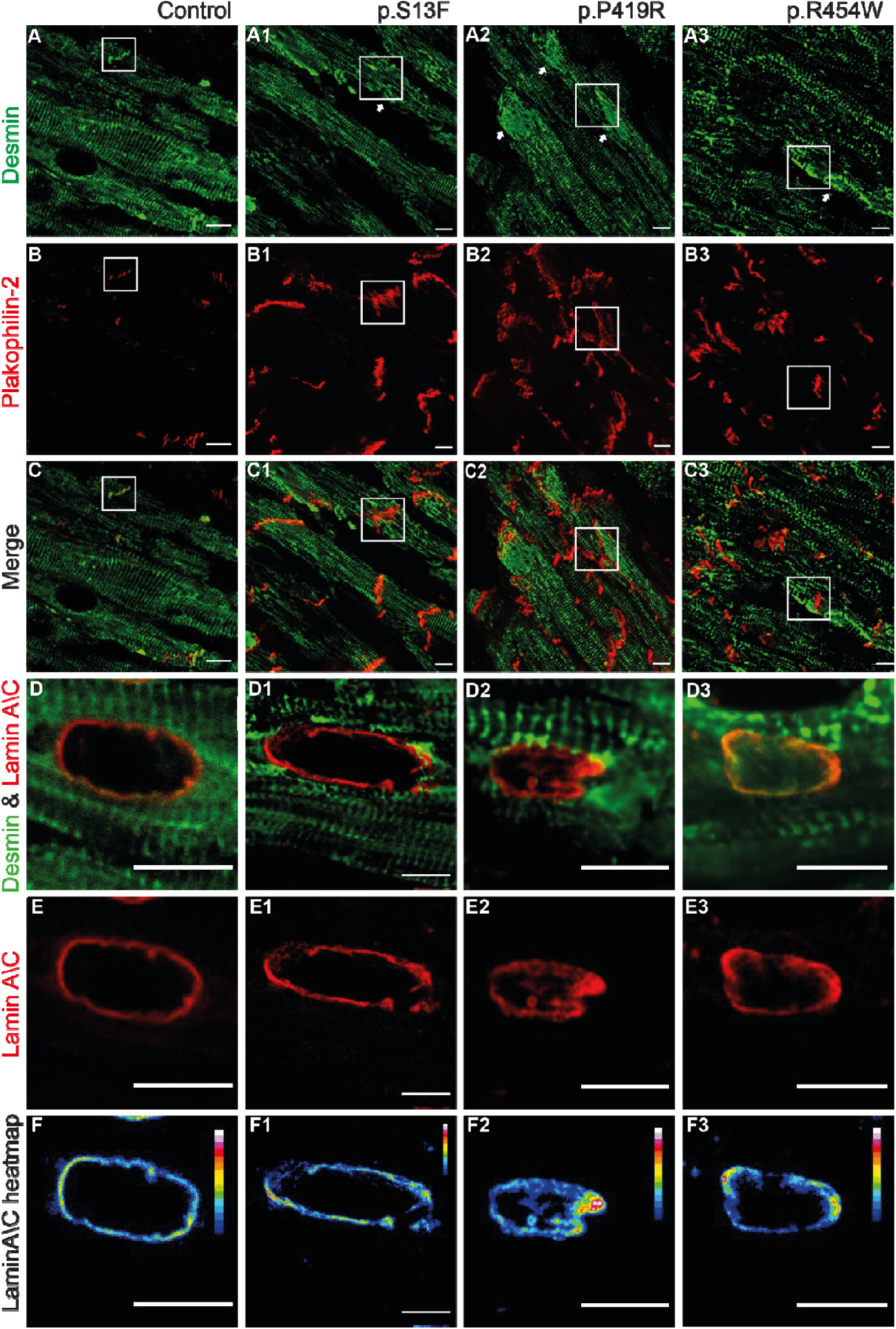
DES variants disrupt the desmin-intercalated disc and perinuclear envelope network in human myocardium. **A–A3**, Representative confocal images of longitudinal human cardiac sections from control myocardium (A) and heterozygous *DES* variant carriers p.S13F (A1), p.P419R (A2), and p.R454W (A3) stained for desmin (green). Control myocardium shows a regular striated desmin pattern with enrichment at IDs. In *DES* variant myocardium, desmin organization is disrupted, with focal desmin aggregates (arrows) and loss or marked reduction of desmin at IDs (boxed regions). **B–B3**, Corresponding plakophilin-2 staining (red). In controls, plakophilin-2 localizes sharply to IDs, whereas *DES* variant myocardium shows clustered and redistributed plakophilin-2 signals at IDs and within the adjacent cytoplasm, frequently in regions with altered desmin organization (boxed regions). **C–C3**, Merged images illustrating the spatial relationship between desmin aggregation, loss of IDs desmin, and plakophilin-2 redistribution, indicating coordinated disruption of the desmin–desmosome/IDs compartment in *DES* variant carriers. **D–D3,** Representative cardiac tissue sections from control myocardium (D) and *DES* variant carriers p.S13F (D1), p.P419R (D2), and p.R454W (D3) showing immunofluorescence for desmin (green) and lamin A/C (red) surrounding individual nuclei. Control tissue shows organized perinuclear desmin with a uniform lamin A/C rim at the nuclear envelope (NE), whereas *DES* variants display perinuclear desmin disorganization and heterogeneous lamin A/C distribution. **E–E3,** Lamin A/C channel alone highlighting altered nuclear lamina organization in *DES* variant myocardium. **F–F3,** Corresponding lamin A/C intensity heat maps (color scale shown) demonstrating polarized lamin A/C distribution at the NE in *DES* variant carriers compared with the uniform nuclear rim observed in controls. White boxes indicate regions of reduced ID-associated desmin. Scale bars, 10 µm.

Beyond cytoplasmic desmin abnormalities, desmin disorganization extended to intercalated discs (IDs). In control myocardium, desmin displayed regular Z-disc striations with enrichment at IDs and colocalization with plakophilin-2. In *DES* variant myocardium, desmin enrichment at IDs was absent (Figure 7C1–C3, white boxes), with redistribution of desmin to cytoplasmic aggregates (arrows). These findings indicate that pathogenic *DES* variants disrupt both the cytoplasmic desmin filament network and its normal association with IDs structures in human myocardium.

Because desmin forms part of the cytoskeletal linkage connecting the sarcomere to the nuclear envelope (NE), we next examined lamin A/C distribution to assess nuclear lamina organization in *DES* variant myocardium Figure 7D-F). Consistent with observations in HL-1 cardiomyocytes, lamin A/C staining frequently showed increased signal intensity at nuclear-envelope regions adjacent to desmin aggregates, whereas nuclear regions lacking nearby aggregates displayed lower lamin A/C intensity (Figure 7F2-F3). This asymmetric lamin A/C distribution suggests localized remodeling of the nuclear lamina associated with perinuclear desmin aggregation.

Together, these findings demonstrate that desmin-network disruption in human myocardium is accompanied not only by cytoplasmic desmin aggregation and loss of desmin organization at IDs, but also by altered nuclear lamina organization, supporting the presence of NE remodeling in *DES* variant carriers.

### 3.8 Desmin-network gene variants define a distinct atrial proteomic remodeling program independent of end-stage heart failure

Because atrial tissue samples from carriers of pathogenic *DES* variants are scarce, variants in functionally related genes (*DES, DSG, PKP2, DSP, and CRYAB)* that converge on the desmin–cytoskeletal–nuclear axis were included, enabling a network-level proteomic analysis. To investigate molecular remodeling associated with DN variants in atrial cardiomyopathy, quantitative proteomic analysis was performed on right atrial (RA) and left atrial (LA) myocardium from DN variant carriers, *TTN* variant carriers, and non-diseased controls.

In RA myocardium, differential expression analysis revealed extensive proteomic remodeling in both disease groups. In the RA DN vs CON comparison, 123 proteins were significantly upregulated and 30 proteins were downregulated, whereas the RA TTN vs CON comparison identified 186 upregulated and 37 downregulated proteins (Figure 8A, B). Because both DN and *TTN* samples were derived from failing hearts, overlap analysis was performed to reduce bias from shared heart-failure–associated remodeling. Venn analysis demonstrated substantial overlap between the DN and *TTN* datasets, including 18 commonly downregulated proteins and 99 commonly upregulated proteins (Figure 8C, D). These overlapping proteins were considered heart-failure–related changes and were therefore excluded from downstream enrichment analyses.

**Figure 8.**
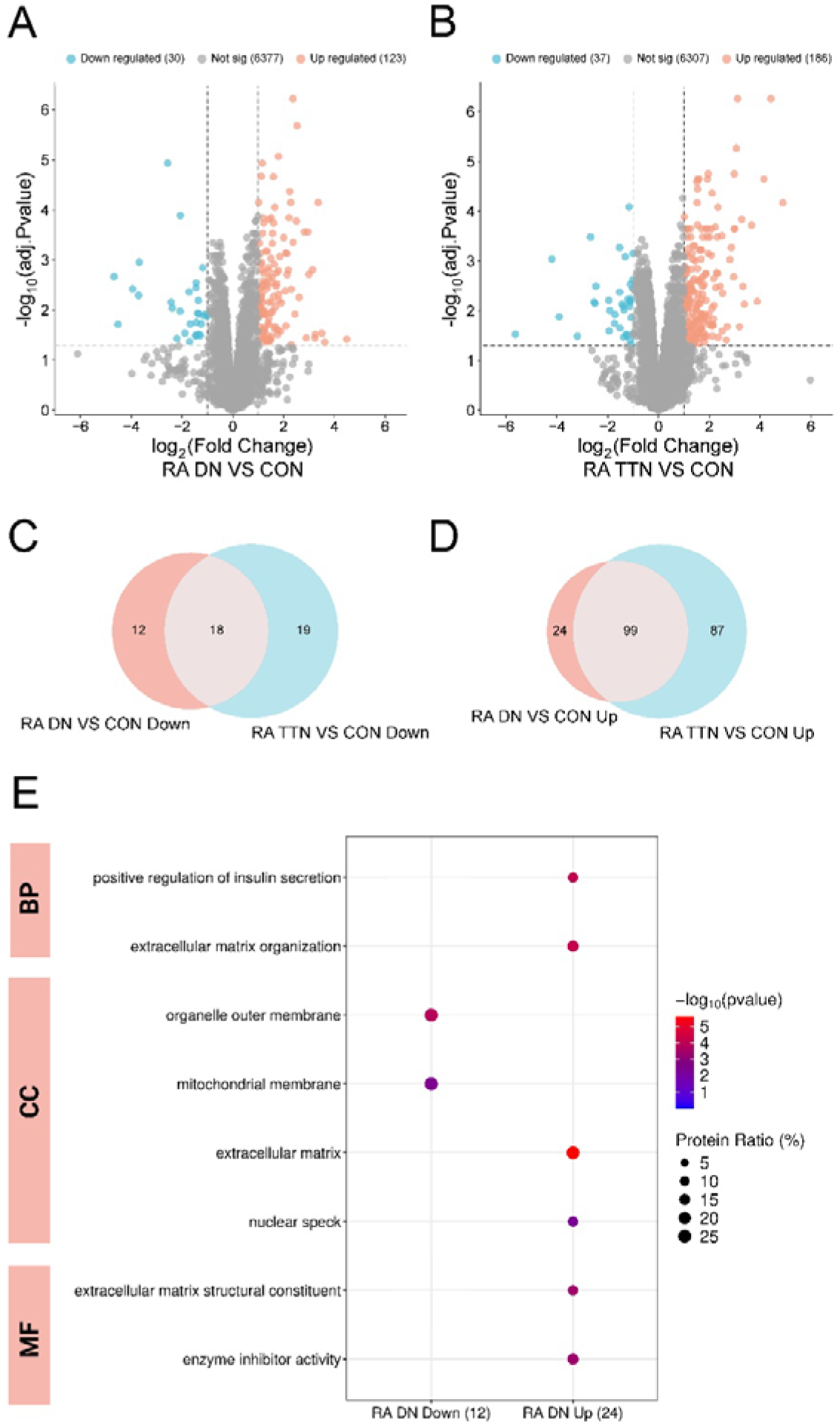
Proteomic remodeling in right atrial myocardium of desmin-network variant carriers. **A**-**B**, Volcano plots showing differentially expressed proteins in RA myocardium from DN variant carriers (A) and *TTN* variant carriers (B) compared with non-diseased controls (CON). Significantly upregulated proteins are shown in red, downregulated proteins in blue, and non-significant proteins in gray. Vertical dashed lines indicate log□ fold-change thresholds and the horizontal dashed line indicates the adjusted *p*-value cutoff. **C-D**, Venn diagrams illustrating overlap of downregulated (C) and upregulated (D) proteins between RA DN vs CON and RA *TTN* vs CON comparisons. Because both DN and *TTN* samples were derived from failing hearts, overlapping proteins were considered shared heart-failure–associated changes and were excluded from downstream enrichment analysis to minimize bias from heart-failure–related remodeling. **E**, Gene Ontology (GO) enrichment analysis of DN-specific proteins remaining after exclusion of proteins overlapping with the RA *TTN* vs CON dataset. Enrichment analysis was performed on 12 uniquely downregulated and 24 uniquely upregulated proteins from the RA DN vs CON comparison. Bubble size represents the proportion of proteins within each GO category and color indicates statistical significance (−log10 *p*-value). Enriched terms are grouped into Biological Process (BP), Cellular Component (CC), and Molecular Function (MF).

GO enrichment analysis of the DN-specific RA protein subsets (12 uniquely downregulated and 24 uniquely upregulated proteins) revealed enrichment of biological processes, including extracellular matrix organization and positive regulation of insulin secretion. Cellular component categories highlighted mitochondrial membrane, organelle outer membrane, extracellular matrix, and nuclear speck compartments, while molecular function analysis revealed enrichment of extracellular matrix structural constituents and enzyme inhibitor activity (Figure 8E).

To further characterize atrial remodeling associated with DN variants, the same analytical workflow was applied to LA myocardium (Supplemental Figure S4). Principal component analysis (PCA) demonstrated separation between CON, DN, and *TTN* samples, indicating distinct global proteomic profiles between groups (Figure S4A–B). Differential expression analysis identified 176 upregulated and 89 downregulated proteins in the LA DN vs CON comparison, whereas the LA *TTN* vs CON comparison revealed 122 upregulated and 68 downregulated proteins (Figure S4C–D).

Overlap analysis demonstrated substantial shared remodeling between DN and *TTN* datasets, consistent with end-stage heart-failure proteomic signatures (Figure S4E–F). After excluding overlapping proteins, GO enrichment analysis of DN-specific proteins revealed pathways related to cytoskeletal organization, supramolecular fiber organization, cell–cell junction organization, and muscle structure development, together with enrichment of mitochondrial organization, extracellular matrix, and actin cytoskeleton cellular components. Molecular function analysis further highlighted extracellular matrix structural constituents, actin binding, protein folding chaperone activity, and enzyme inhibitor activity (Figure S4G).

Together, integration of RA and LA proteomic datasets indicate that DN variants are associated with a distinct atrial remodeling program involving cytoskeletal and intermediate-filament–associated pathways, nuclear-related compartments, extracellular matrix remodeling, mitochondrial organization, and proteostasis-related chaperone activity, supporting a desmin-dependent atrial remodeling program driving cardiomyopathy.

## 4. Discussion

In this study, we identify a mechanistic framework linking pathogenic *DES* variants to atrial cardiomyopathy. Using HL-1 atrial cardiomyocytes and validation in human cardiac tissue, we show that aggregation-prone variants, particularly p.N342D and p.S13F, disrupt the perinuclear desmin scaffold, destabilize the LINC complex, and remodel the nuclear lamina with focal lamin A/C polarization. These structural alterations are accompanied by increased cGAS-positive nuclear foci and electrophysiological remodeling characterized by shortened action potential duration and impaired Ca²⁺ transients. Importantly, atrial proteomic profiling of DN variant carriers revealed enrichment of pathways related to cytoskeletal organization, chaperone-mediated proteostasis, extracellular matrix signaling, and nuclear-associated compartments, providing tissue-level support for the nuclear injury phenotype observed in desmin variant–expressing atrial cardiomyocytes.

### 4.1 Desmin aggregation remodels nuclear architecture

A central finding of our study is that pathogenic *DES* variants remodel nuclear architecture in atrial cardiomyocytes, particularly when desmin forms perinuclear aggregates, as observed for p.S13F and p.N342D. Previous work demonstrated that the desmin–nuclear linkage is required for nuclear homeostasis in cardiomyocytes, as disruption of desmin tethering perturbs nuclear structure and cardiomyocyte function.^27^ This linkage is mediated in part through nesprin-3, which connects intermediate filaments to the outer nuclear membrane via plectin. Consistent with this model, WT and p.R454W largely preserved a filamentous DN and showed a uniform lamin A/C rim, whereas p.S13F and p.N342D formed perinuclear aggregates associated with polarized lamin A/C enrichment. The spatial coupling between aggregates and lamina remodeling suggests localized mechanical loading of the NE rather than a global stress response. Lamin A/C is known to reorganize in response to mechanical forces, supporting the concept that intermediate filament networks regulate nuclear mechanics.^27^ Recent work further demonstrated dynamic interactions between desmin, plectin, and lamin A/C at the nuclear periphery under mechanical stress.^28^ Our observation of nesprin-3 disorganization in aggregation-prone variants extends this framework and links variant-specific desmin aggregation to disruption of the desmin–plectin–nesprin axis in atrial cardiomyocytes.

### 4.2 DES variants induce nuclear envelope instability

Disruption of the DN was also associated with NE instability. Cardiomyocytes expressing p.N342D and p.S13F exhibited a marked increase in cGAS-positive foci near the NE, consistent with exposure of nuclear DNA to the cytoplasm. cGAS accumulation is widely used as a marker of NE rupture or loss of nuclear compartmentalization in mechanically stressed cells.^14^ NE rupture has been extensively described in laminopathies and NE cardiomyopathies, where impaired lamina integrity leads to chromatin herniation and DNA leakage.^13^ Our data extend these observations by identifying DN disruption as an upstream cause of NE stress in atrial cardiomyocytes. Importantly, nuclear lamina polarization, nesprin-3 disorganization, and cGAS accumulation were observed both in HL-1 cardiomyocytes and in myocardium from *DES* variant carriers, supporting a conserved mechanism in which perinuclear desmin aggregation disrupts nucleocytoskeletal coupling and predisposes the NE to mechanical failure. Although cGAS foci do not quantify rupture dynamics or downstream STING signaling, their spatial association with nuclear deformation supports a model of structurally induced nuclear injury.^29^

### 4.3 Variant location determines nuclear phenotype

Our findings also highlight the importance of variant location within the desmin protein. Desmin consists of an N-terminal head domain, a central rod domain, and a C-terminal tail domain, each contributing differently to filament assembly and network stability.^30^ Prior studies showed that head- and rod-domain variants frequently disrupt filament assembly and induce cytoplasmic aggregation, whereas tail-domain variants more often preserve filament formation while altering network mechanics.^31^ Consistent with these genotype–structure relationships, the head-domain variant p.S13F and rod-domain variant p.N342D showed pronounced perinuclear aggregation and nuclear remodeling, whereas the tail-domain variant p.R454W largely preserved filament organization. Aggregation-prone variants also exhibited the strongest lamina remodeling, nesprin-3 disruption, and cGAS accumulation, suggesting that variant-dependent aggregation determines nuclear stress severity. These observations support the concept that pathogenic *DES* variants act through a toxic gain-of-function mechanism, in which misassembled desmin perturbs intracellular architecture and mechanically loads specific cellular compartments.^26,32^

### 4.4 Desmin-network disruption alters cardiomyocyte function

Structural abnormalities were accompanied by electrophysiological remodeling, including shortened APD□□ and APD□□, reduced Ca²⁺ transient amplitude, and diminished Δ-voltage fluorescence amplitude.^33–35^ Shortened action potential duration reduces atrial refractoriness and promotes re-entrant activity, whereas impaired Ca²⁺ cycling can facilitate triggered activity.^36,37^ Notably, the magnitude of electrical remodeling correlated with structural severity, with p.N342D cardiomyocytes exhibiting the strongest aggregation and electrophysiological phenotype. These findings support the concept that cytoskeletal and nuclear integrity are closely linked to atrial electrophysiology. Previous work has shown that perturbation of nuclear–cytoskeletal coupling alters ion-channel expression and Ca²⁺ handling in cardiomyocytes.27

At the tissue level, atrial proteomics of DN variant carriers revealed enrichment of pathways related to extracellular matrix organization, mitochondrial compartments, chaperones, and nuclear-associated domains after subtraction of *TTN*-associated end-stage heart-failure signatures. These alterations parallel the cellular phenotypes observed in HL-1 cardiomyocytes, including desmin aggregation, nuclear envelope remodeling, and activation of chaperone pathways. The concordance between the cellular model and human atrial proteomics supports the relevance of desmin-driven cytoskeletal and nuclear remodeling in atrial cardiomyopathy.

### 4.5 HSPs induction mitigates desmin-mediated injury

Pharmacological induction of HSPs using GGA reduced desmin aggregation, improved nuclear morphology, and partially normalized electrophysiological abnormalities, particularly in p.N342D-expressing cardiomyocytes. HSPs regulate protein quality control and cytoskeletal stability, and their protective role in atrial remodeling and experimental AF has been demonstrated previously.^2,33^ Our findings extend these observations to genetic atrial disease and support the concept that impaired proteostasis contributes to desmin aggregation and downstream remodeling. Although our data provide proof-of-principle in a cellular model, they do not address long-term efficacy or arrhythmia suppression *in vivo*. The ongoing GENIALITY trial is currently evaluating the effects of GGA in postoperative AF.^38^ Results from this study will provide important insights into the translational potential of proteostasis-targeting strategies for atrial disease.

### 4.6 Limitations

Several limitations should be acknowledged. HL-1 cardiomyocytes, while atrial-derived, do not fully recapitulate the electrophysiological complexity of adult human atrial cardiomyocytes, although this model has successfully identified multiple druggable pathways.^33,34,39,40^ cGAS accumulation indicates nuclear envelope instability but does not define rupture kinetics or downstream signaling. Finally, availability of human myocardial tissue was limited and proteomic analyses relied on network-based grouping of desmin-associated variants.

### 4.7 Conclusions

Pathogenic *DES* and DN gene variants cause a desmin-dependent atrial cardiomyopathy characterized by perinuclear cytoskeletal aggregation, nuclear envelope instability with cGAS accumulation, and electrophysiological remodeling. Integration of cellular, electrophysiological, proteomic, and human tissue data identifies nuclear–cytoskeletal uncoupling as a central disease mechanism and highlights proteostasis enhancement as a potentially modifiable pathway. These findings expand the molecular framework of inherited atrial cardiomyopathy and provide a mechanistic basis for arrhythmic risk associated with DN dysfunction.

## Data Availability

The data that support the findings of this study are available from the corresponding authors upon reasonable request.

## Nonstandard Abbreviations and Acronyms

AF: Atrial fibrillation
APD: Action potential duration
APD_20_: /APD_90_ Action potential duration at 20% / 90% repolarization
CaT: Calcium transients
cGAS: Cyclic GMP–AMP synthase
DIA-MS: Data-independent acquisition mass spectrometry
DN: Desmin-network
DMSO: Dimethyl sulfoxide
FBS: Fetal bovine serum
FDR: False discovery rate
GGA: Geranylgeranylacetone
GO: Gene ontology
GO-BP / GO-CC / GO-MF: Gene ontology: Biological process / Cellular component / Molecular function
HSP(s): Heat shock protein(s)
IDs: Intercalated discs
KEGG: Kyoto encyclopedia of genes and genomes
LA / RA: Left atrium / right atrium
LINC: Linker of nucleoskeleton and cytoskeleton
NE: Nuclear envelope
PCA: Principal component analysis
PFA: Paraformaldehyde
RT: Room temperature
SEM: Standard error of the mean
WT: Wild-type

